# *CYP2D6* Genotyping for Personalized Therapy of Tamoxifen in Indonesian Women with ER+ Breast Cancer

**DOI:** 10.1101/2021.06.25.21259564

**Authors:** Baitha Palanggatan Maggadani, Kathleen Irena Junusmin, Levana L. Sani, Caroline Mahendra, Margareta Amelia, Gabriella, Astrid Irwanto, Harmita, Yahdiana Harahap, Samuel J. Haryono

**Author notes:** These authors contributed equally to this work (Baitha Palanggatan Maggadani, Kathleen Irena Junusmin).

## Abstract

Tamoxifen is a Selective Estrogen-Receptor Modulator (SERM) commonly prescribed for standard of care in estrogen receptor positive (ER+) breast cancer as an adjuvant therapy. Tamoxifen is metabolized by *CYP2D6* into its active metabolite, endoxifen, which has been known to play an important role in reducing risk of ER+ breast cancer recurrence. *CYP2D6* is a highly polymorphic gene with more than 100 alleles. The phenotype of this gene is categorized into ultrarapid metabolizer (UM), normal metabolizer (NM), intermediate metabolizer (IM), and poor metabolizer (PM). Certain *CYP2D6* polymorphisms may cause reduced activity of this enzyme. Studies have found that reduced *CYP2D6* activity in IM and PM patients causes low efficacy of standard tamoxifen therapy. This study aims to observe the distribution of *CYP2D6* alleles and its correlation with endoxifen levels in Indonesian ER+ breast cancer patients. 151 patients who have received tamoxifen therapy for at least eight weeks were recruited prospectively. DNA and blood samples were collected with buccal swab and finger-prick methods, respectively. Genotyping was performed using the qPCR method while metabolite level measurement was performed using high performance liquid chromatography tandem mass spectrometry. We found that 40.67% of ER+ breast cancer patients recruited were IM. *CYP2D6*10* was the most abundant allele (0.288) in this population, and *\*10/*36* was the most frequently observed diplotype (0.236). Endoxifen levels between the NM-PM, NM-IM, and IM-PM were statistically significant (*p*-value = 6.26 × 10^−5^, 9.12 × 10^−5^, and 4.714 × 10^−3^, respectively), and dose increase of tamoxifen to 40 mg daily successfully increased endoxifen levels in IMs to a similar level with NMs at baseline. Given these findings, implementing pharmacogenomic testing of *CYP2D6* on ER+ breast cancer women who are about to undergo tamoxifen therapy may be beneficial to increase the likelihood of achieving expected endoxifen levels, thus better treatment efficacy.

## Introduction

Estrogen receptor (ER) expression is the main indicator of potential responses to hormonal therapy, and approximately 70% of human breast cancers are hormone-dependent and ER+ (Lumachi *et al*., 2013). Hormone receptor–positive BC is associated with less aggressive features and a better prognosis because of the benefits from currently available endocrine therapy (Li *et al*., 2020). Tamoxifen is the current standard of care for ER+ breast cancer adjuvant therapy. It works by binding to the estrogen receptor. The drug has been proven effective in reducing the number of recurrences especially in pre-menopausal women. About 170,000 tamoxifen prescriptions were filed in 2015 in Indonesia (IMS prescription data Indonesia, 2015), which implies that the usage of this drug has been prevalent in Indonesia to treat ER+ breast cancer.

Tamoxifen is a prodrug that needs to be metabolized to be active. However, half of the patients receiving tamoxifen may not have the full benefit of this drug due to the genetic polymorphisms that affect the function of the main enzyme metabolizing tamoxifen, CYP2D6 (Goetz *et al*., 2008). Tamoxifen is metabolized to 4-hydroxy-N-desmethyltamoxifen (endoxifen), which has been proven to be an important contributor to the overall anticancer effect. Endoxifen is formed predominantly by CYP2D6 from N-desmethyltamoxifen, the most abundant metabolite (Dezentjé *et al*., 2009). Endoxifen threshold value has been discovered to significantly impact breast cancer survival rates (Goetz *et al*., 2005; Madlensky *et al*., 2011). Upon years of follow up, those with endoxifen levels lower than 5.97 ng/mL had a 30% higher chance of having recurrence of breast cancer. Madlensky *et al*. further showed that being a CYP2D6 poor/intermediate metabolizer was associated with having a higher Body Mass Index (BMI), and consequently lower tamoxifen concentrations predicted risk for breast cancer recurrence (Madlensky *et al*., 2011). Additionally, study has also shown that individual variability of *CYP2D6* contributed 53% towards the ratio of N-desmethyltamoxifen and endoxifen, while combined other CYPs genetic factors (*CYP2C9, CYP2C19, CYP3A5*) and non-genetic factors (age, BMI) contributed to only 2.8% (Saladores *et al*., 2015).

*CYP2D6* gene that encodes Cytochrome P450 2D6 (CYP2D6) enzyme has more than 100 variants; some causing reduced activity, and others causing complete loss of function. The spectrum of the *CYP2D6* enzymatic activity translates to different metabolizer profiles that are grouped into normal, ultrarapid, extensive, intermediate, and poor metabolizers (NM, UM, EM, IM, and PM, respectively), depending on how many reducing and/or loss of function alleles an individual carries. Asians and Africans were known to have up to 50% reduced activity alleles (Bradford, 2002). In Malays, Chinese and Indians, intermediate metabolizers occur in 35%, 45.38%, and 15%, respectively (Teh *et al*., 2001; Cui *et al*., 2020; Goh *et al*., 2017). Meanwhile, Caucasians were commonly extensive metabolizers (Chin *et al*., 2016). *CYP2D6* ultra-rapid and extensive metabolizers are able to take tamoxifen as indicated, according to the guidelines by Clinical Pharmacogenetics Implementation Consortium (CPIC) (Goetz *et al*., 2018).

This study aims to observe the distribution of *CYP2D6* genotypes and its correlation with endoxifen levels in ER+ breast cancer patients in Indonesia. *CYP2D6* allele frequency and tamoxifen metabolite concentrations were observed. Patients who had *CYP2D6* IM and PM phenotype profile were given recommendation to adjust tamoxifen dose to 40 mg daily, while patients who were clinically ineligible for tamoxifen dose increase according to clinical guidelines were switched to aromatase inhibitor. Our study observed the effectiveness of adjusting tamoxifen dosage as the first line of action for patients who are clinically eligible to still consume the drug. Patients who received tamoxifen dose adjustment were monitored to ensure safety from potential side effects associated with tamoxifen.

## Materials and Methods

### Ethics Approval

Institutional Review Board (IRB) approval was granted by MRCCC Siloam Hospitals Semanggi Ethics Review Committee (Jakarta, Indonesia) under IRB Reference Number 001/EA/KEPKK/RSMRCCC/V/2019.

### Study Participants

Patients were recruited from SJH Initiative, MRCCC Siloam Hospital Jakarta, Indonesia, from October 2019 to April 2021 (n=151). The inclusion criteria of this study were as follows: (1) patient was diagnosed with ER+ breast cancer and (2) had consumed tamoxifen for at least eight weeks. Patients who fulfilled the inclusion criteria were offered to participate in the study and informed consent was obtained. Flow of recruitment steps is shown in Fig. 1. Ethnicities reported in this study were self-reported, participants who identified with two or more ethnicities were categorized as mixed races.

**Fig. 1.**
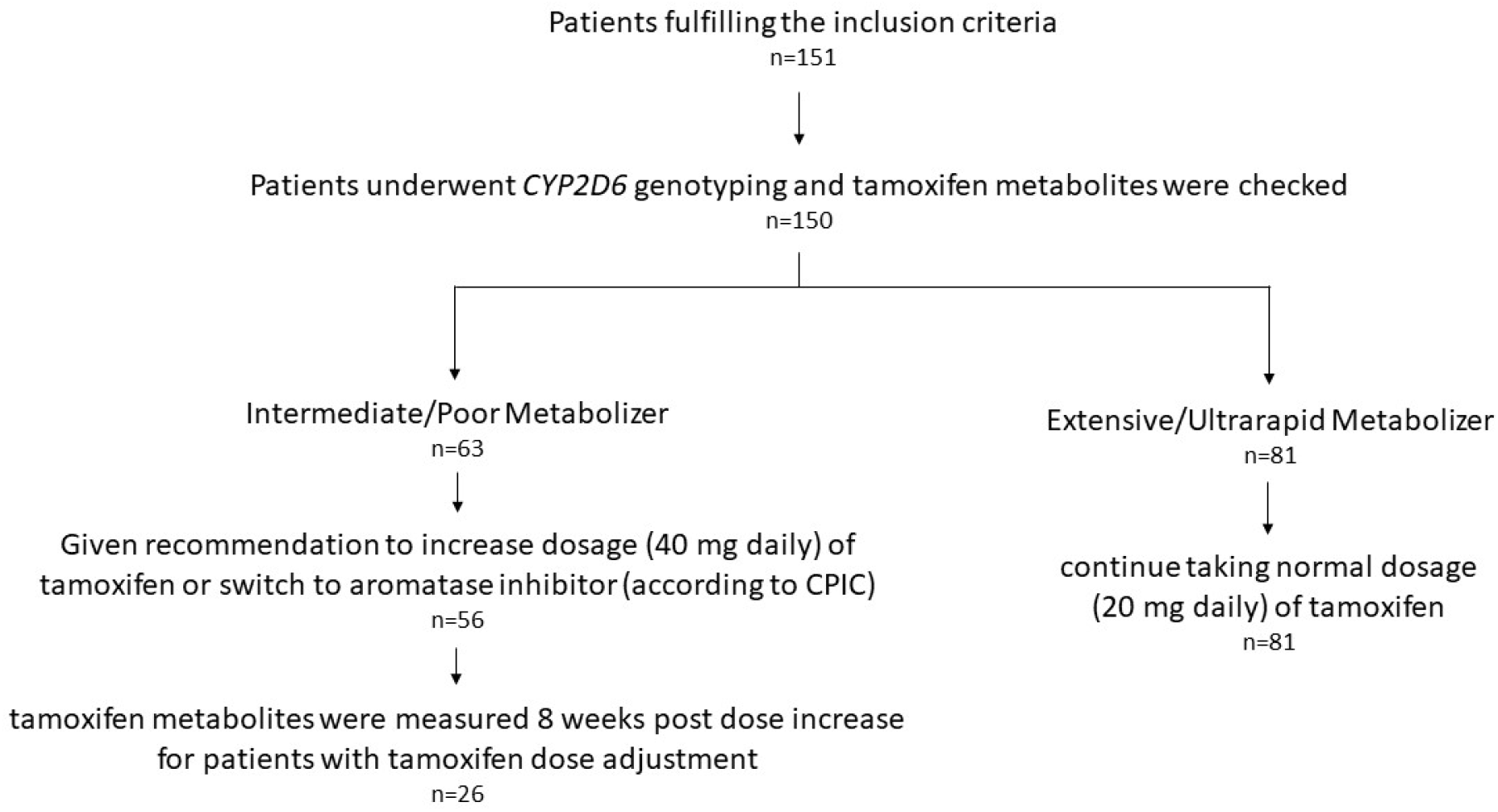
Research flow diagram.

### DNA Extraction

Buccal swab sample was obtained from the patient for *CYP2D6* genotyping using ORAcollect•DNA OCR-100 (DNA Genotek) swab. Genomic DNA were extracted from buccal swab samples using Monarch Genomic DNA Purification Kit (NEB #T3010) following the manufacturer’s instructions. Concentration of gDNA extracts were quantified using BioDrop spectrophotometer. Acceptance criteria to further process the DNA extract for genotyping, include: (1) total DNA yield ≥ 500 ng, (2) A260/280 ratio ≥ 1.75, and (3) A260/230 ratio ≥ 1.75.

### *CYP2D6* Genotyping

*CYP2D6* genotyping was performed using Nala PGx Core™, a Lab-Developed Test genotyping panel consisting of four pharmacogenes: *CYP2D6, CYP2C19, CYP2C9* and *SLCO1B1* (Kothary *et al*., 2021). *CYP2D6* variants that were genotyped in this test included rs35742686, rs59421388, rs3892097, rs5030656, rs72549352, rs5030655, rs28371725, rs16947, rs1065852, rs267608319, rs769258, rs5030865, rs1135840, total copy number of intron 2 and a detection for the presence of exon 9 conversion. Genomic DNA extracts were diluted to 2ng/uL and added as template for Nala PGx Core™ qPCR runs on Bio-Rad CFX96 Touch™ Real-Time PCR Detection System. *CYP2D6* haplotypes, diplotypes and phenotypes were inferred by Nala Clinical Decision Support™ which is a class A medical device (Health Sciences Authority, Singapore) compatible with Nala PGx Core™ qPCR output.

### Measurement of Tamoxifen Metabolites

Finger-prick blood sample was obtained using Volumetric Absorptive Microsampling (VAMS) technique. VAMS extraction was performed in methanol by sonication-assisted extraction method for 25 minutes after 2 hours of VAMS drying. Separation was carried out using Acquity UPLC BEH C_18_ column (2.1 × 100 mm; 1.7 µm), with a flow rate of 0.2 mL/minute, and the mobile phase gradient of formic acid 0.1% combined with formic acid 0.1% in acetonitrile for 5 minutes. The UPLC-MS/MS Waters Xevo TQD Triple Quadrupole with MassLynx Software controller (Waters, Milford, USA) was employed in metabolites measurement. Mass detection was carried out utilizing Triple Quadrupole (TQD) with Multiple Reaction Monitoring (MRM) analysis modes and an electrospray ionization source using positive mode. The method was developed in the Bioavailability and Bioequivalence Laboratory of Universitas Indonesia and validated according to FDA and EMA guidelines (Maggadani *et al*., 2021). The multiple reaction monitoring (MRM) value were set at m/z 372.28>72.22 for TAM; 374.29>58.22 for END; 388.29>72.19 for 4-HT; 358.22>58.09 for NDT; and 260.20>116.20 for propranolol as the internal standard.

### Patient Follow Up

Patients with IM or PM *CYP2D6* profile who were clinically ineligible for tamoxifen dose increase were switched to aromatase inhibitor (n=18) and were not followed up further for side effects monitoring and metabolite levels changes. This group of patients were determined based on clinical judgement according to the available guidelines by The National Surgical Oncologist Organization and Ministry of Health in Indonesia (Komite Penanggulangan Kanker Nasional, n.d.), National Comprehensive Cancer Network (NCCN, 2021), and British Columbia Cancer Agency (Kennecke *et al*., 2006). IM or PM patients who did not have any contraindications to tamoxifen were given a recommendation to adjust its dose to 40 mg/day (n=26), while UMs and NMs remained with the normal 20 mg/day recommended dose (n=81). Tamoxifen metabolites levels in the study participants who were given 40 mg/day of tamoxifen were measured eight weeks post dose adjustment. Endocrine symptoms which were possible side effects of tamoxifen therapy were also monitored in patients who received tamoxifen dose adjustment to 40 mg daily using the FACT-ES questionnaire (Fallowfield *et al*., 1999).

### Data Analysis

Data and statistical analysis were performed using Microsoft® Excel® for Microsoft 365 and R version 4.0.3. Deviation from Hardy-Weinberg equilibrium was performed on the haplotype frequencies using the chi-square statistical test, where Bonferonni correction was applied to determine the *p*-value threshold for significant deviation. Analysis of Variance (ANOVA) test was used to see if metabolite levels distribution at baseline were statistically different across all metabolites, followed by a paired T-test between each pair of metabolites when significance was found. Distribution of metabolite levels before and after dose adjustment was compared using a T-test, and the same test was used to compare the distribution of metabolite levels in IMs post-dose adjustment against NMs (baseline). Concerning symptoms related to endocrine therapy post-dose adjustment on IMs were compared against NMs. Chi-square test was performed per symptom to check for the difference between the two groups.

## Results

### Demographics of Study Participants

Table 1 shows that out of the 151 participants included in the study, most of the participants were 50 years old and below, making up 78.15% of the total respondents. This proportion was followed by participants between 51-59 years old (17.88%). A small number of older participants with age ≥60 years (3.97%) was also observed. The majority of participants consisted of individuals with Chinese (33.77%) and Javanese (25.17%) descents. Participants with multiethnic and multiracial descents were also observed (16.56%), followed by small numbers of other Indonesian ethnicities such as Sundanese (5.96%), Batak (5.3%), Betawi (3.31%), Minang (3.31%), Ambonese (1.32%), and South Sumatran (1.32%). Among these participants, 47.33% underwent lumpectomy (also known as breast conserving surgery), while 44% underwent mastectomy (total removal of breast tissue). Aside from surgical intervention, 66.67% of these participants underwent adjuvant post-operative radiotherapy and 50% underwent adjuvant chemotherapy. Respondents were mostly still in the early stage of breast cancer during the time of recruitment, with proportion as follows: stage I (27.15%), stage IIa (23.84%), and stage IIb (13.91%). Participants who were enrolled to the study and were in the later stage of breast cancer were also observed, with proportion as follows: stage IIIa (7.95%), IIIb (5.96%), and stage IV (7.95%). About half of the study participants (50.33%) were enrolled within 12 months after initial diagnosis of breast cancer. The other participants were enrolled within 13-24 (15.23%), 25-36 (13.25%), and 37-48 (9.27%) months after initial diagnosis, with a proportion of patients who had been diagnosed for longer than four years ago (10.6%). According to the available biopsy data, 44.37% of the participants had moderately differentiated tumors, while 27.81% and 11.92% of the participants had poorly and moderately differentiated tumors, respectively.

**Table 1.**
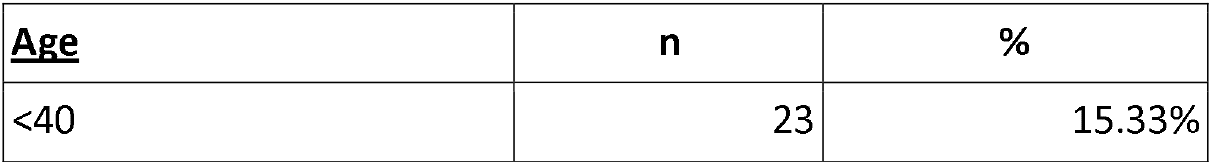
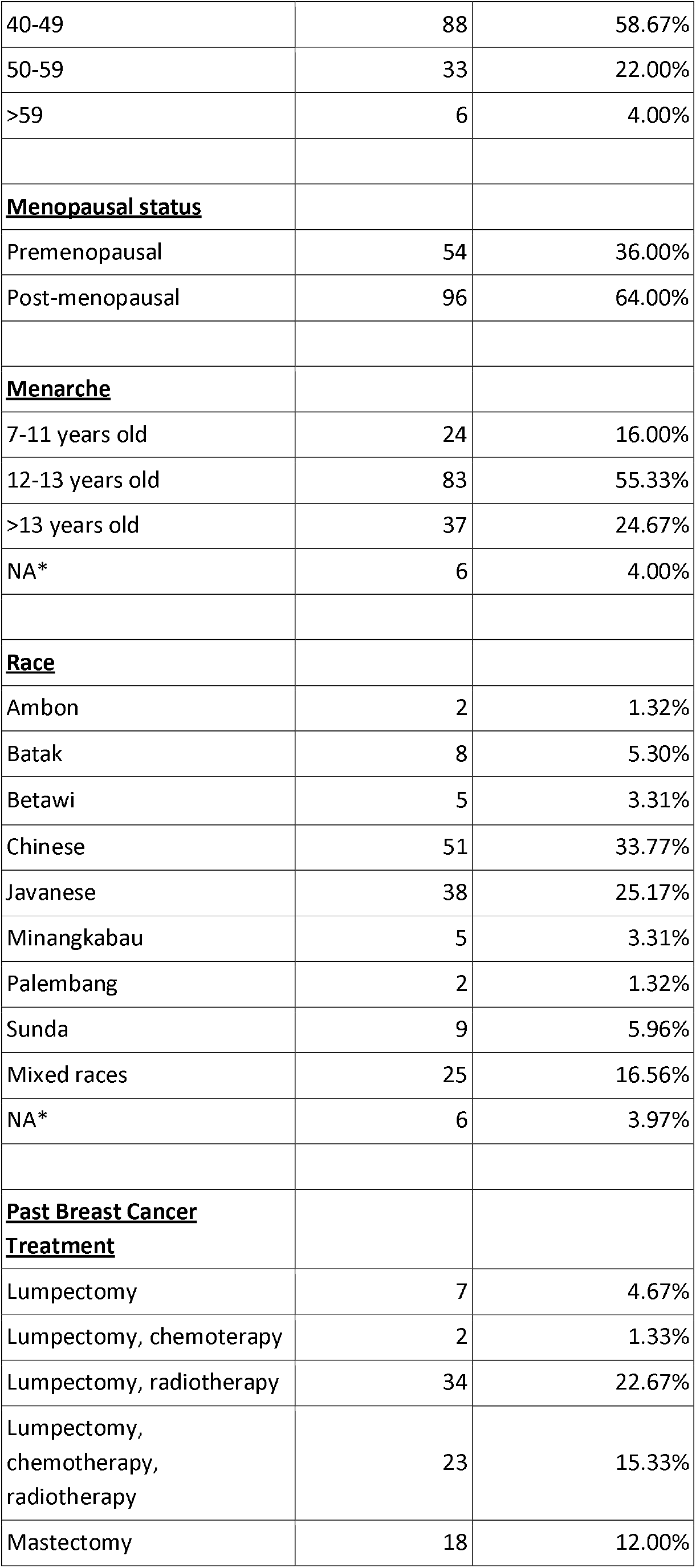
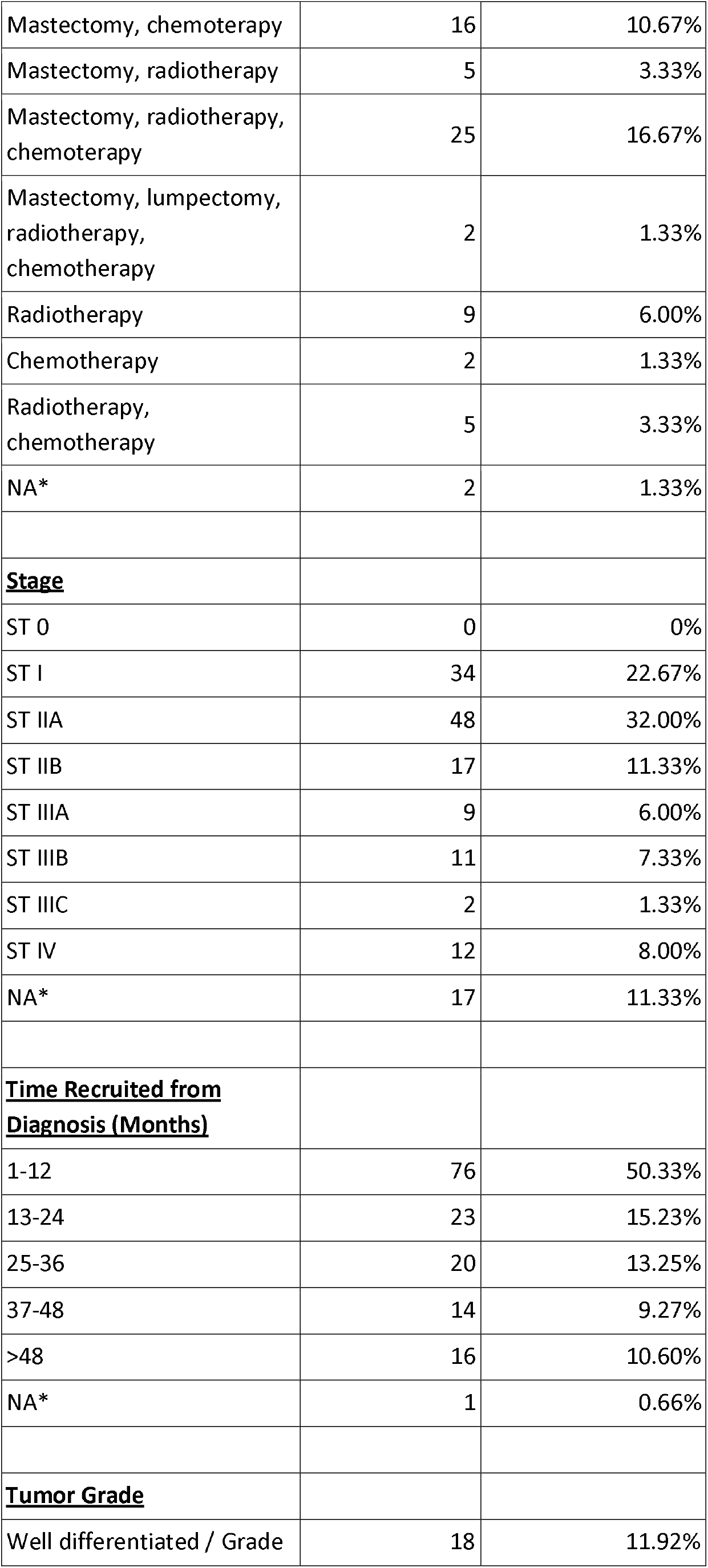
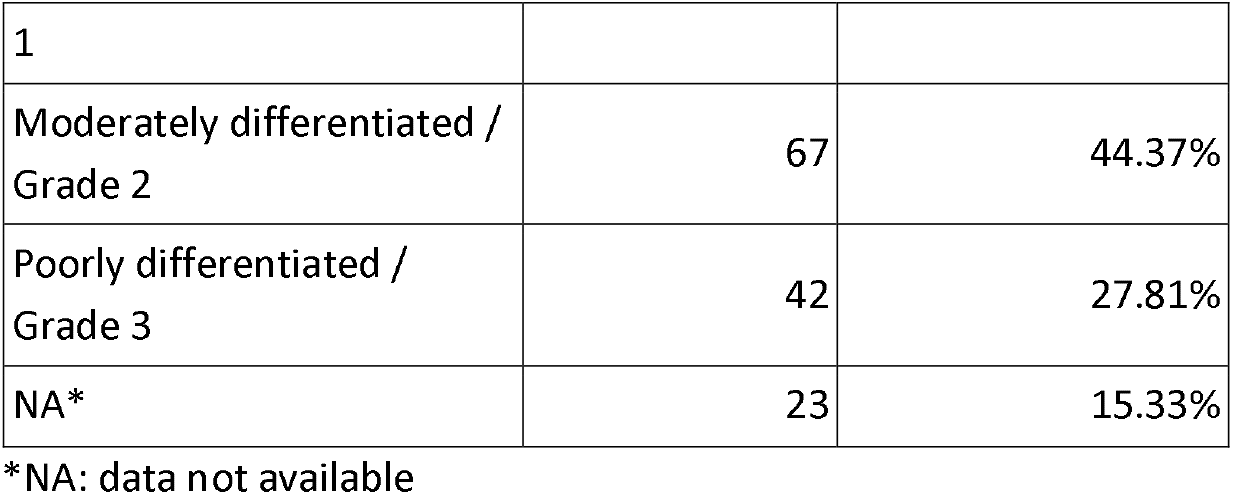
Study respondents demographics.

### *CYP2D6* Haplotype Distribution

All haplotypes observed were in Hardy-Weinberg equilibrium (*p*-value > 0.005). *CYP2D6*10* was found to be the most abundant haplotype in the population (0.288, n=83/288), followed by *CYP2D6*36* (0.253, n=73/288). Compared to PharmGKB database of the East Asian population, *\*10* was lower, but *\*36* was much higher in this study compared to the frequency reported by the database, 0.012 (Fig. 2). The reference haplotype *CYP2D6*1* was observed with frequency of 0.233 (n=67/288), and other haplotypes were also observed with frequencies as follows: *\*2* (0.128, n=37/288), *\*41* (0.045, n=13/288), *\*5* (0.021, n=6/288), *\*3* (0.014, n=4/288), *\*39* (0.007, n=2/288), *\*4A* (0.007, n=2/288), and *\*14* (0.003, n=1/288).

**Fig. 2.**
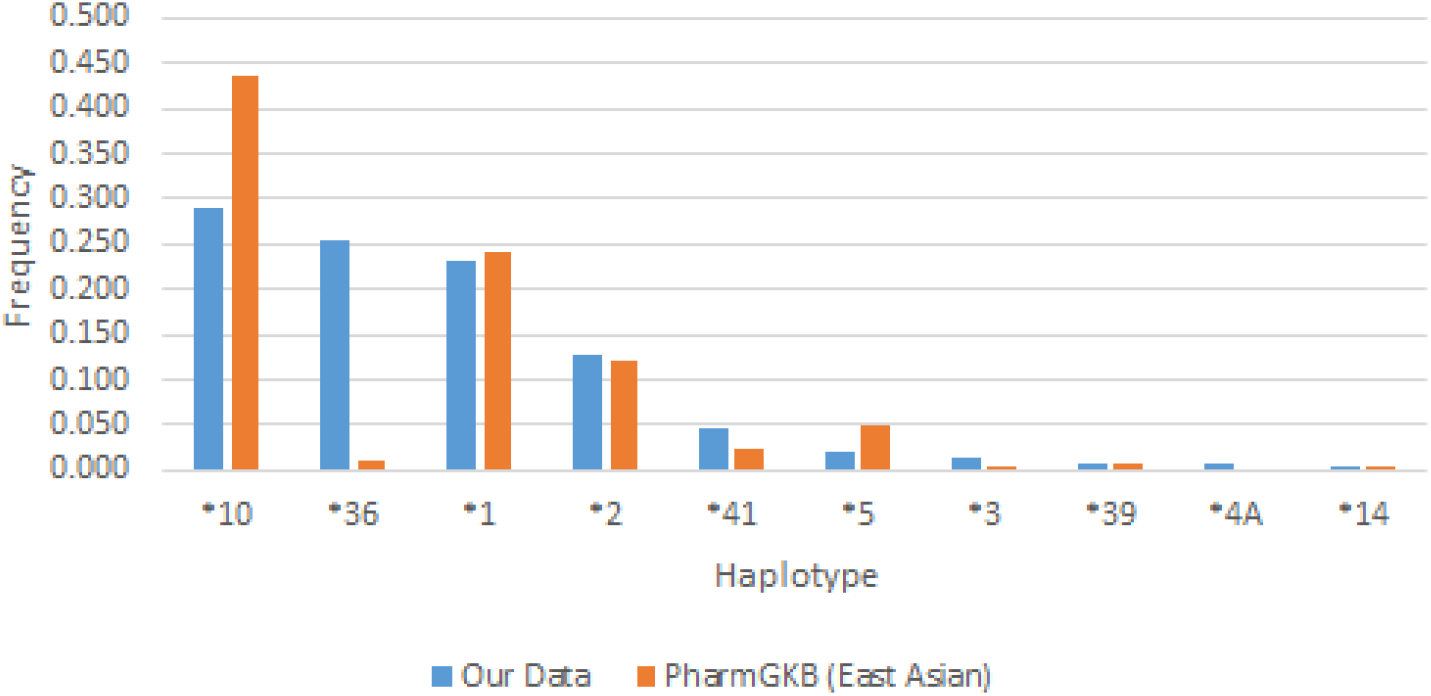
Distribution of haplotype frequencies among Indonesian breast cancer patients. n=288

### *CYP2D6* Diplotype Distribution

Our study demonstrated *\*10/*36* (0.236, n=34/144) as the most abundant diplotype in the population, followed by *\*1/*36* (0.132, n=19/144) (Table 2). Other diplotypes that were observed in this study with diplotype frequencies between 0.1-0.05 were as follows: *\*2/*10* (0.097, n=14/144), *\*1/*1* (0.09, n=13/144), *\*2/*36* (0.083, n=12/144), *\*1/*10* (0.076, n=11/144), and *\*10/*10* (0.065, n=9/144). Other diplotypes observed had frequencies lower than 0.05. The list of relevant diplotypes can be found in Table 2.

**Table 2.** *CYP2D6* diplotype frequencies observed.

| Diplotype | Phenotype | Counts (N total = 144) | Frequency |
| --- | --- | --- | --- |
| *10/*36 | Intermediate Metabolizer | 34 | 0.236 |
| *1/*36 | Normal Metabolizer | 19 | 0.132 |
| *2/*10 | Normal Metabolizer | 14 | 0.097 |
| *1/*1 | Normal Metabolizer | 13 | 0.090 |
| *2/*36 | Normal Metabolizer | 12 | 0.083 |
| *1/*10 | Normal Metabolizer | 11 | 0.076 |
| *10/*10 | Normal Metabolizer | 9 | 0.065 |
| Others^ |  | 41 | 0.222 |

^Other diplotypes were observed with frequency less than 0.05, these diplotypes were \*1/\*2, \*36/\*41, \*1/\*41, \*10/\*41, \*1/\*5, \*2/\*2, \*3/\*36, \*5/\*10, \*5/\*41, \*1/\*3, \*1/\*4A, \*14/\*36, \*2/\*3, \*2/\*39, \*2/\*41, \*36/\*39, and \*4A/\*10

### *CYP2D6* Phenotypes Distribution

Our findings show that among the 150 patients genotyped, 40.67% (n=61/150) were IMs. This is much higher than the current known global prevalence of IMs which is between 0.4-11%. The frequency of NMs observed in this study was 54% (n=81/150). PMs were also observed in the population at 1.33% (n=61/150) (Fig. 3). Ultrarapid metabolizers were not observed among the participants in this study. Distribution of the *CYP2D6* phenotypes among major ethnicities in this study’s participants showed a higher proportion of IMs in Chinese (56.86%, n=29/51) compared to other ethnicities such as Javanese (23.68%, n=9/38). PM was observed in the Javanese group with 2.63% frequency (n=1). Ethnicities with participant counts less than 10 were grouped as others, due to inefficient number of samples to conclude allele frequencies (Supplementary Table 2). Mixed races group showed 37.5% proportion of IM (n=6/16). Among all major ethnicity groups, only Chinese ethnicity group displayed a greater proportion of IM compared to NM (Fig. 4).

**Fig. 3.**
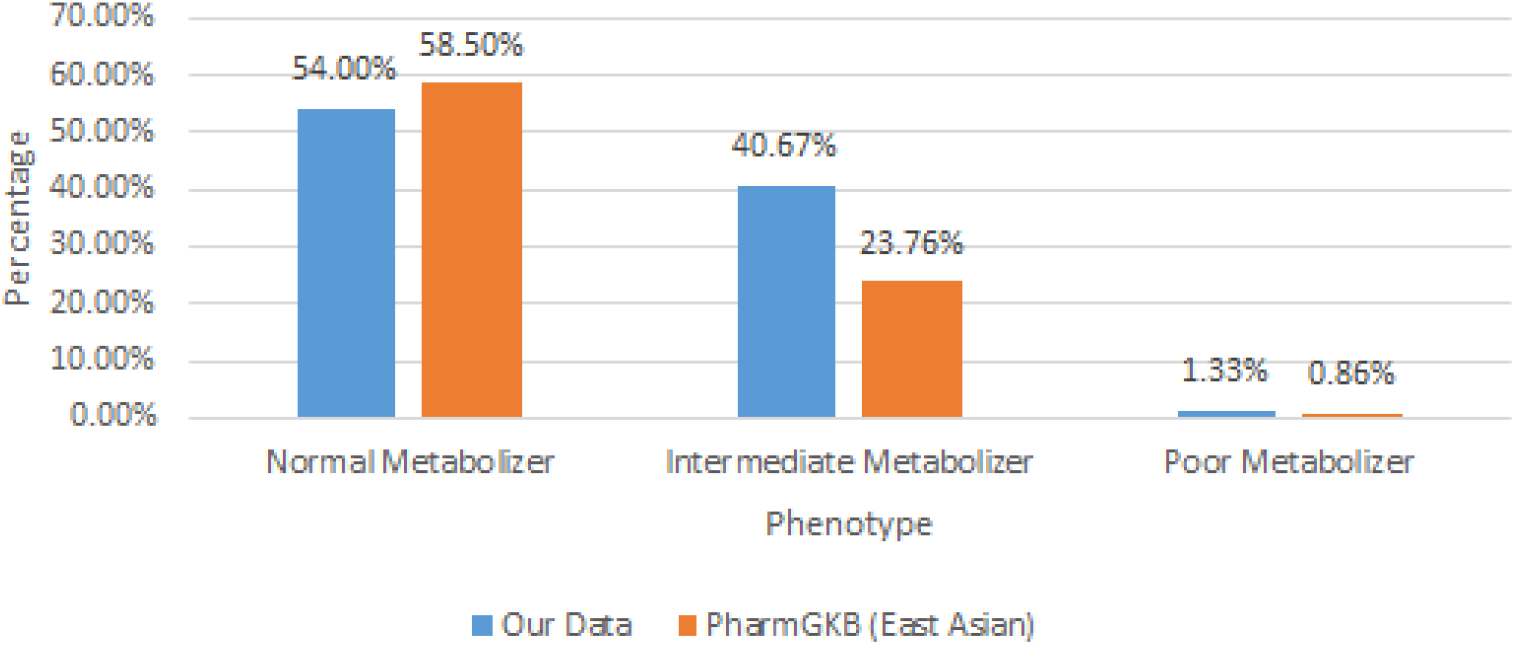
Distribution of phenotype frequencies among Indonesian breast cancer patients. n=144

**Fig. 4.**
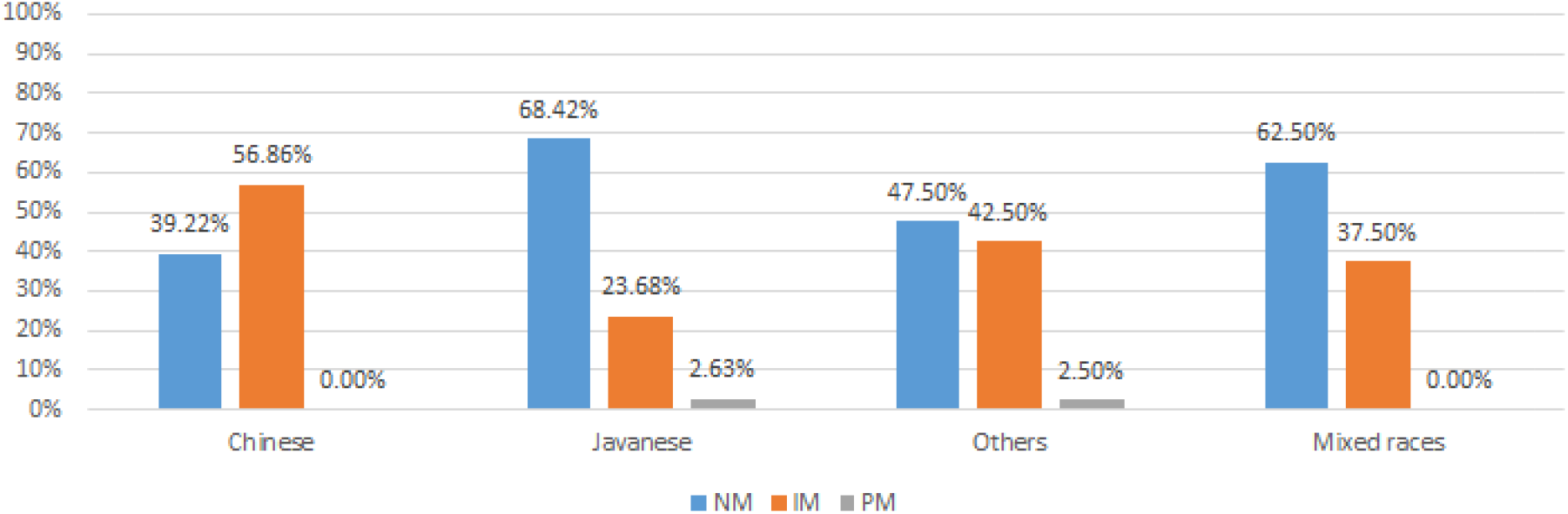
Distribution of phenotype frequencies per major ethnicity among Indonesian breast cancer patients. n=151

### Tamoxifen Metabolite Concentration

Endoxifen levels among the three metabolizers were significantly different (*p*-value = 0.00307, Table 3). The rest of the metabolites did not show any statistically significant distribution among phenotypes (*p*-value = 0.964, 0.461, 0.443 for tamoxifen, 4-hydroxytamoxifen, and N-desmethyltamoxifen, respectively). T-test performed on endoxifen levels for each phenotype pair displayed significant difference among all phenotype pairs (*p*-value = 6.26 × 10^−5^, 9.12 × 10^−5^, and 4.714 × 10^−3^ for NM-PM, NM-IM, and IM-PM, respectively), demonstrating distinction of endoxifen levels across different phenotypes (Fig. 5). After grouping the endoxifen levels into five quintiles, it was revealed that the highest number of IMs fall into the lowest quintile while the highest number of NMs fall into the highest quintile (Supplementary Table 1).

**Table 3.** Summary of metabolite levels in relation to *CYP2D6* metabolizer profiles.

| *CYP2D6*<br>Phenotype |  | Concentration (ng/mL) |  |  |  |
| --- | --- | --- | --- | --- | --- |
|  |  | Tamoxifen | Endoxifen | 4OH-tam | ND-tam |
| Normal<br>Metabolizer<br>(N = 81) | Mean | 82.30 | 13.17 | 3.30 | 224.86 |
|  | SD | 35.21 | 6.62 | 1.46 | 56.83 |
|  | Median | 77.46 | 11.98 | 3.07 | 240.59 |
|  | Range | 31.22 - 170.82 | 3.55 - 34.77 | 1.5 - 7.66 | 80.63 - 321.88 |
| Intermediate Metabolizer (N = 61) | Mean | 85.62 | 9.59 | 3.60 | 233.70 |
|  | SD | 37.20 | 4.35 | 1.67 | 58.01 |
|  | Median | 81.72 | 8.33 | 3.27 | 241.55 |
|  | Range | 14.22 - 210.39 | 3.17 - 22.97 | 1.5 - 9.31 | 77.61 - 337.29 |
| Poor Metabolizer (N = 2) | Mean | 91.49 | 4.52 | 3.24 | 276.45 |
|  | SD | 33.93 | 0.83 | 0.26 | 90.44 |
|  | Median | 91.49 | 4.52 | 3.24 | 276.45 |
|  | Range | 67.49 - 115.48 | 3.94 - 5.11 | 3.06 - 3.43 | 212.5 - 340.41 |
| *p*-value (ANOVA) |  | 0.964 | 0.00307* | 0.461 | 0.443 |

**Fig. 5.**
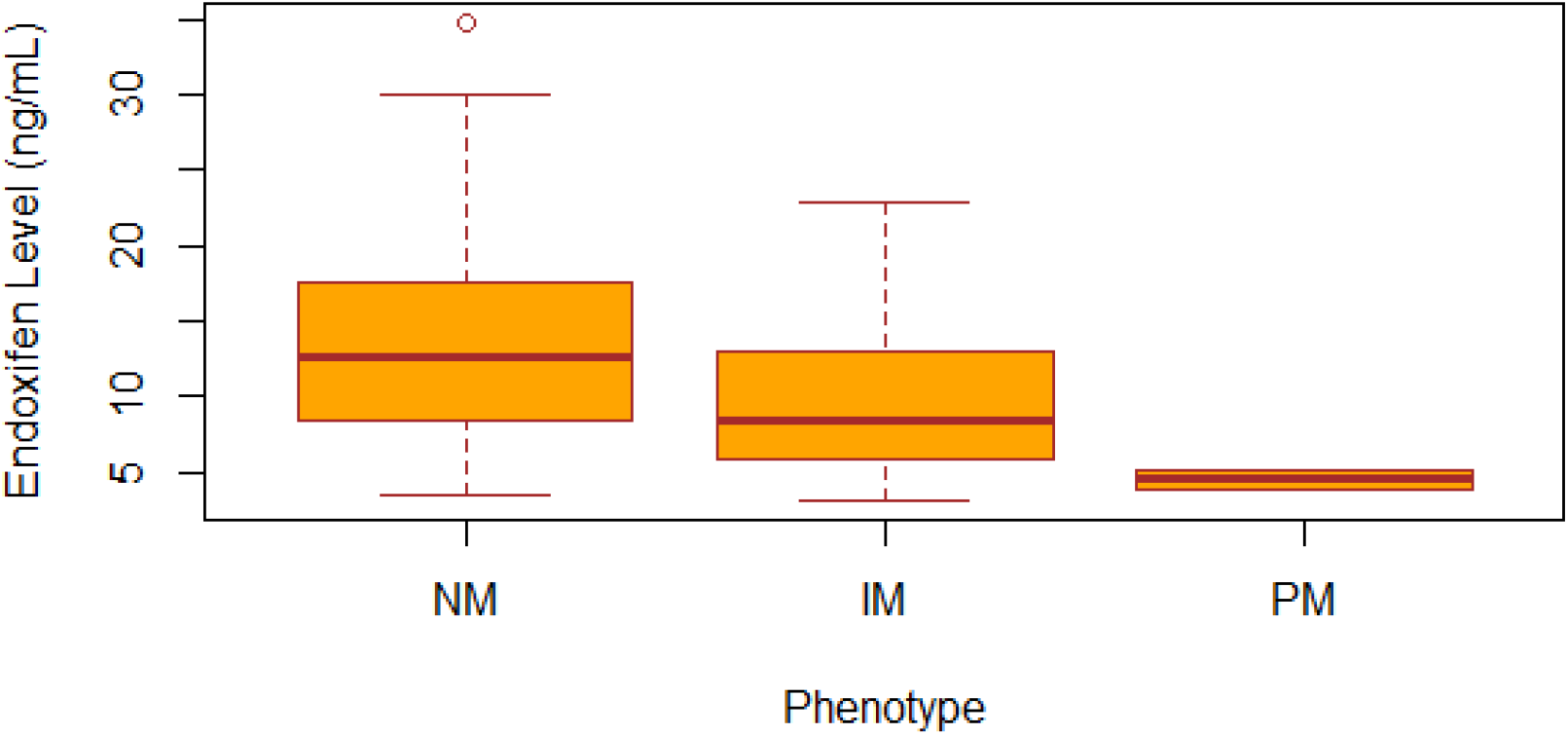
Distribution of endoxifen levels for each observed phenotype at the baseline. Normal metabolizer/NM (n=81), Intermediate metabolizer/IM (n=61), Poor Metabolizer/PM (n=2)

### Follow Up Action Following PGx Testing

Among 66 IM or PM participants who were given the recommendation to modify their medication based on their *CYP2D6* phenotype (Fig. 6), 18 patients (27.3%, n=18/66) had their medication switched to aromatase inhibitors based on clinical guidelines or certain medical procedure such as post Ovarian Function Suppression (OFS) endocrine therapy. 38 patients (57.6%, n=38/66) were recommended by their physicians to adjust their tamoxifen dosage from 20 mg daily to 40 mg daily, while the remaining participants who did not follow the genotype-guided recommendation either passed away or experienced recurrence, thus they had to dismiss their adjuvant therapy temporarily (15.2%, n=10/66).

**Fig. 6.**
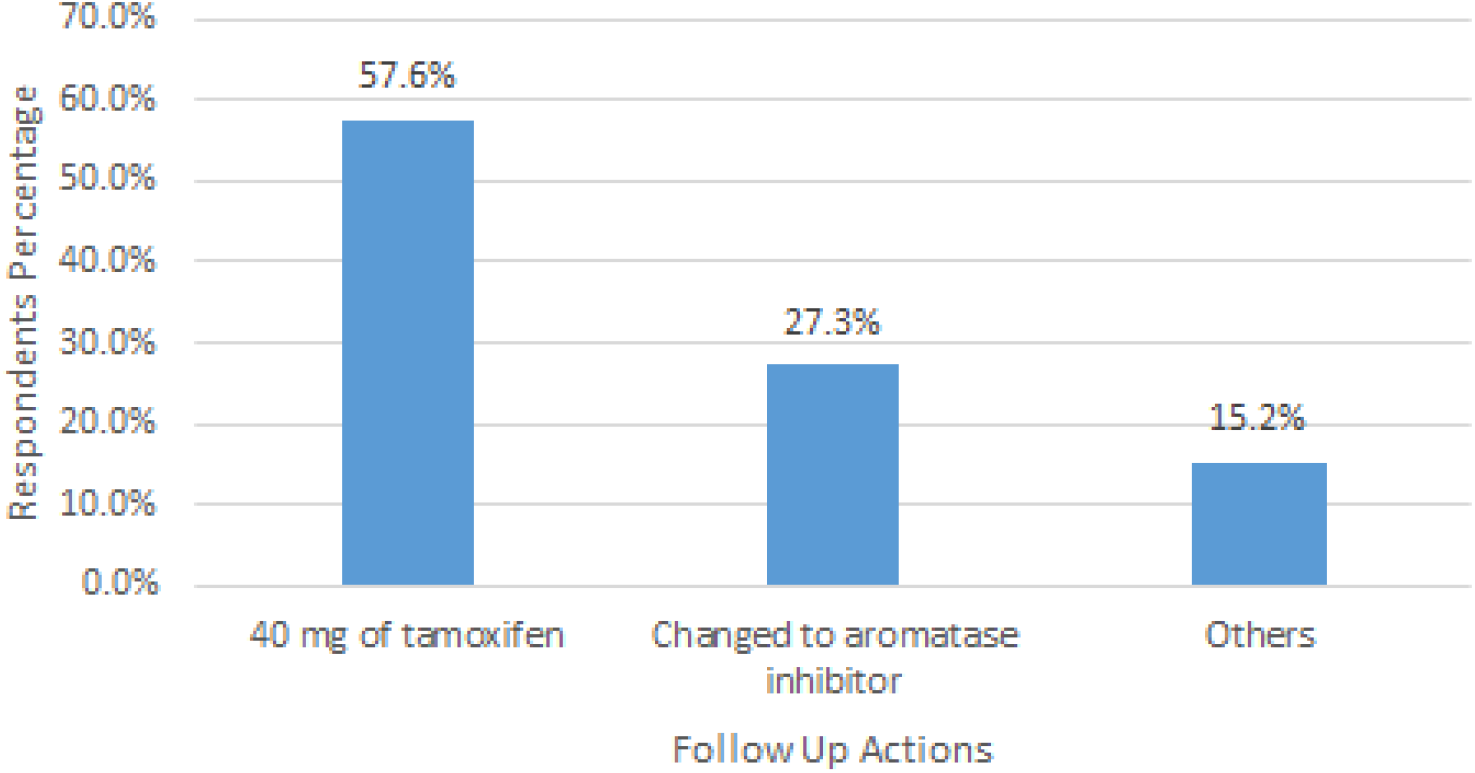
Distribution of the different follow up actions selected by doctors after patient’s *CYP2D6* profile was characterized through genetic testing. n=66

### Metabolite Levels Post Dose Adjustment

26 patients who took 40 mg of tamoxifen daily for two months all experienced an increase in metabolite levels. After dose adjustment, the range of tamoxifen metabolites increased as follows: tamoxifen levels from 14.22-210.39 ng/mL to 80.59-254.96 ng/mL; endoxifen levels from 3.17-22.97 ng/mL to 7.68-23.36 ng/mL; 4-hydroxytamoxifen levels from 1.5-9.31 ng/mL to 3.34-12.99 ng/mL, and N-desmethyltamoxifen levels from 77.61-337.29 ng/mL to 236.8-501.9 ng/mL (Fig. 7). Metabolite levels before and after dose adjustment had *p*-value < 0.05, demonstrating statistically significant differences before and after dose adjustment across all metabolites.

**Fig. 7.**
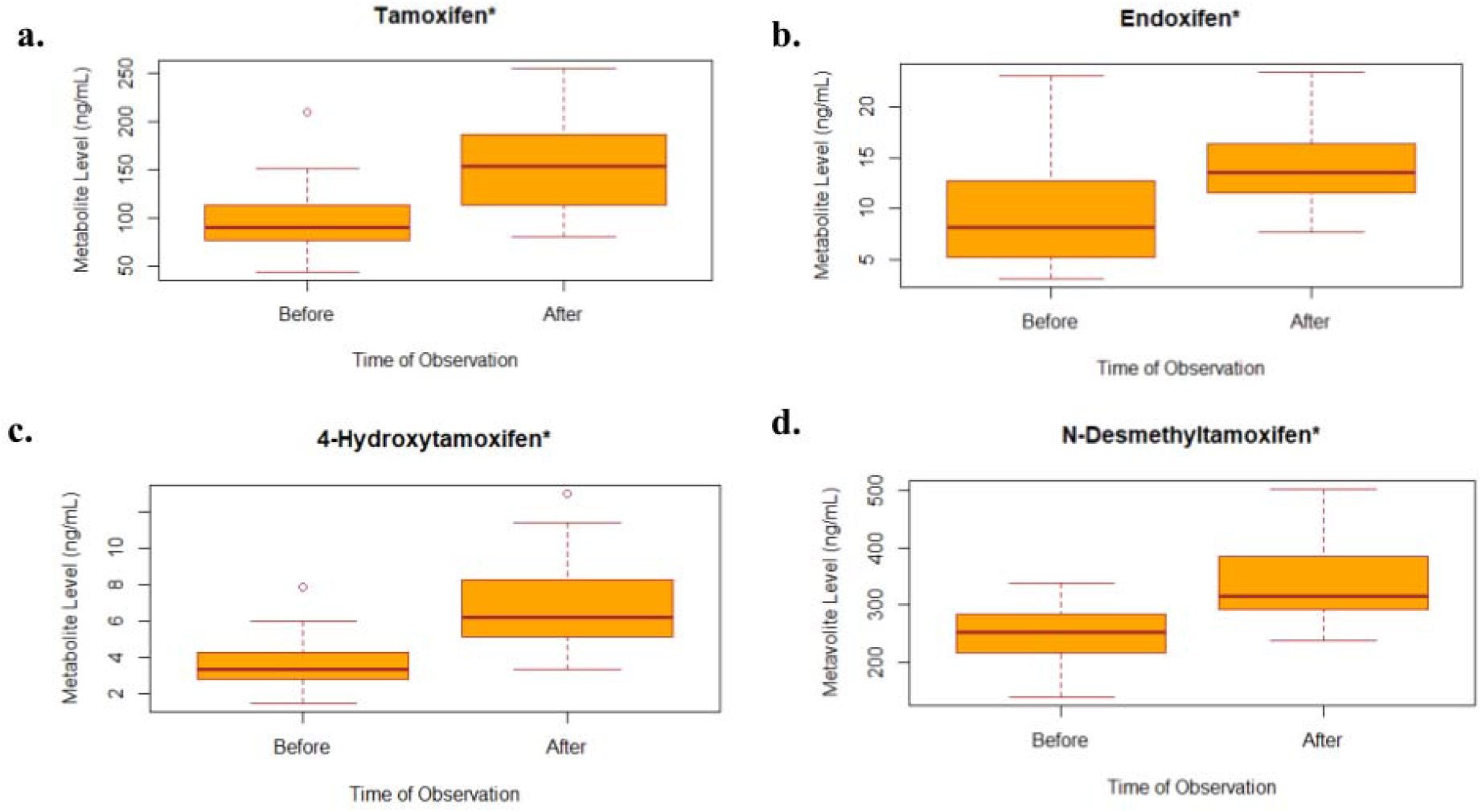
Metabolite levels before and after dose adjustment for IM patients. a.) Tamoxifen, b.) endoxifen, c.) 4-hydroxytamoxifen, d.) N-desmethyltamoxifen. *Statistically significant *p*-values were observed between metabolites before and after dose adjustment, n=26

The metabolite levels in IMs (n=26) post dose adjustment were compared against NMs (n=81) as the baseline, showing indeed a significant difference between the two groups (*p*-value < 0.05) for all metabolites except endoxifen (*p*-value = 0.4135). The distribution of endoxifen levels in IMs post dose adjustment (7.68-23.36 ng/mL) were similar to the endoxifen levels in NMs (3.55 - 34.77 ng/mL) at baseline (Fig. 8).

**Fig. 8.**
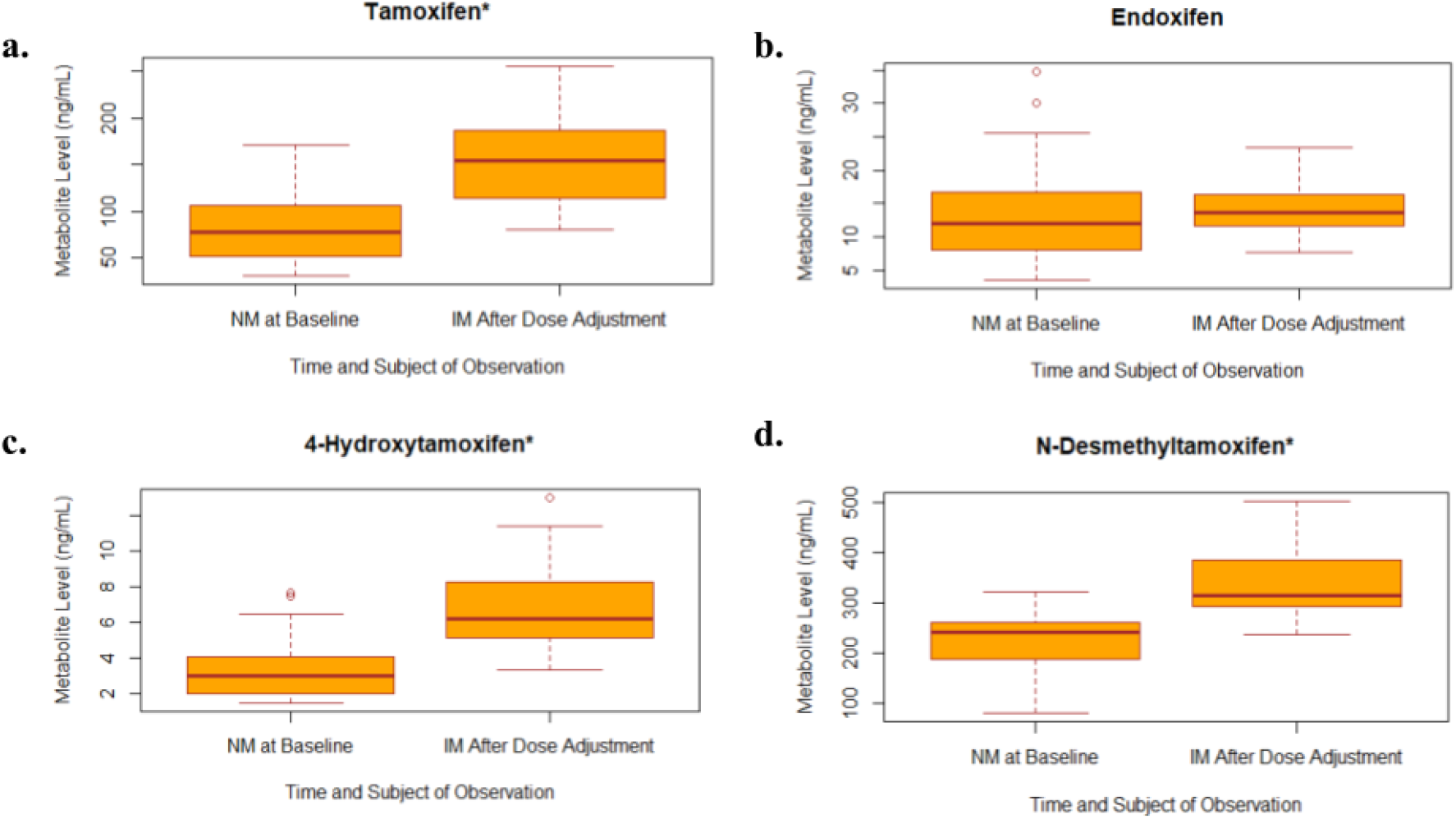
Metabolite levels in IMs after dose adjustment compared to NMs at the baseline. a.) Tamoxifen, b.) endoxifen, c.) 4-hydroxytamoxifen, d.) N-desmethyltamoxifen. *Statistically significant *p*-values were observed, n=81 (NMs), n=26 (IMs). Endoxifen levels in IMs post dose adjustment were statistically similar to NMs at the baseline.

### Side Effects Post Dose Adjustment

The most commonly reported treatment side effects in IMs were weight gain and mood swings, which are related to endocrine therapy. These occurred in 65.83% of participants who received 40 mg of tamoxifen daily (n=17/26). Other common symptoms related to hormonal changes were also observed in participants who received 40 mg of tamoxifen daily such as hotflush (50%, n=13/26), cold sweats (19.23%, n=5/26), night sweats (26.92%, n=7/26), vaginal discharge (42.31%, n=11/26), vaginal itching or irritation (15.38%, n=4/26), vaginal bleeding or spotting (23.08%, n=6/26), vaginal dryness (11.54%, n=3/26), pain or discomfort during intercourse (3.85%, n=1/26), lost interest in sex (15.38%, n=4/26), breast sensitivity or tenderness (53.85%, n=14/26), and irritability (61.54%, n=16/26). Other symptoms that might be related to endocrine therapy were also observed, such as lightheaded/dizziness (34.62%, n=9/26), vomiting (3.85%, n=1/26), headaches (53.85%, n=14/26), bloating (46.15%, n=12/26), and pain in joints (50%, n=13/26). No post-dose adjustment participants reported diarrhea.

The most commonly reported side effect in the patient group that took 20 mg of tamoxifen daily was mood swings, occuring in 74.19% of the respondents (n=23/31), although they did not receive any treatment adjustments. Other common symptoms related to hormonal changes were also observed in NM participants such has hotflush (35.48%, n=11/31), cold sweats (12.9%, n=4/31), night sweats (29.03%, n=9/31), vaginal discharge (38.71%, n=12/31), vaginal itching or irritation (22.58%, n=7/31), vaginal bleeding or spotting (16.13%, n=5/31), vaginal dryness (32.26%, n=10/31), pain or discomfort during intercourse (51.61%, n=16/31), lost interest in sex (64.52%, n=20/31), breast sensitivity or tenderness (41.94%, n=13/31), and irritability (58.06%, n=18/31). Other symptoms that might be related to endocrine therapy were also observed, such as lightheaded/dizziness (35.48%, n=11/31), vomiting (6.45%, n=2/31), diarrhea (3.23%, n=1/31), headaches (29.03%, n=9/31), bloating (38.71%, n=12/31), and pain in joints 67.74%, n=21/31).

T-test performed between symptoms experienced by participants receiving dose adjustment to 40 mg daily and participants taking 20 mg daily resulted in two symptoms (pain or discomfort during intercourse and lost interest in sex) with statistical significance between the two groups. Other than these two symptoms, the other symptoms did not have significant difference among the two groups, indicating that dose escalation up to 40 mg daily did not increase potential toxicity or side effects (Table 4). Thrombophlebitis, thrombosis, endometriosis, and endometrial cancer were also some of the most concerning side effects of tamoxifen (American Society of Clinical Oncology, 2009; Bergman *et al*., 2000), and none of these side effects were observed in the observed population.

**Table 4.** Number and percentage of patient responses related to adverse events in FACT-ES post eight weeks after dose adjustment. *Statistically significant *p*-value was observed

| Symptoms | NM participants who received 20 mg of tamoxifen daily (N = 31) |  | IM participants who received 40 mg of tamoxifen daily (N = 22) |  | *p*-value |
| --- | --- | --- | --- | --- | --- |
|  | Patients reported side effect (n) | Patients reported side effect (%) | Patients reported side effect (n) | Patients reported side effect (%) |  |
| Hot Flushes | 11 | 35.48% | 13 | 50.00% | 0.269361 |
| Cold Sweats | 4 | 12.90% | 5 | 19.23% | 0.717648 |
| Night sweats | 9 | 29.03% | 7 | 26.92% | 0.86249 |
| Vaginal discharge | 12 | 38.71% | 11 | 42.31% | 0.777297 |
| Vaginal itching/irritation | 7 | 22.58% | 4 | 15.38% | 0.492987 |
| Vaginal bleeding or spotting | 5 | 16.13% | 6 | 23.08% | 0.507122 |
| Vaginal dryness | 10 | 32.26% | 3 | 11.54% | 0.063252 |
| Pain or discomfort with intercourse* | 16 | 51.61% | 1 | 3.85% | $8.48 \times 10^{-5}$* |
| Lost interest in sex | 20 | 64.52% | 4 | 15.38% | 0.005461* |
| Weight gain | 20 | 64.52% | 17 | 65.38% | 1 |
| Lightheaded (dizzy) | 11 | 35.48% | 9 | 34.62% | 1 |
| Vomiting | 2 | 6.45% | 1 | 3.85% | 1 |
| Diarrhea | 1 | 3.23% | 0 | 0.00% | 1 |
| Headaches | 9 | 29.03% | 14 | 53.85% | 0.057089 |
| Bloating | 12 | 38.71% | 12 | 46.15% | 0.571608 |
| Breast sensitivity/tenderness | 13 | 41.94% | 14 | 53.85% | 0.371093 |
| Mood swings | 23 | 74.19% | 17 | 65.38% | 0.470842 |
| Irritable | 18 | 58.06% | 16 | 61.54% | 0.791337 |
| Pain in joints | 21 | 67.74% | 13 | 50.00% | 0.173783 |

\*Statistically significant *p*-values were observed between IMs who have received tamoxifen dose adjustment and NMs who took the standard dose

## Discussion

This study aimed to observe the distribution of *CYP2D6* genotypes and phenotypes across Indonesian women diagnosed with ER+ breast cancer who were taking tamoxifen as adjuvant therapy. Our respondents were mostly of Chinese and Javanese descent. Chinese ethnicity group in this study’s population showed a higher proportion of intermediate metabolizers, while the Javanese ethnicity group was dominated by normal metabolizers (Fig. 4). The proportion of IMs in Indonesian Chinese included in this study was higher than a similar study conducted on Han Chinese population, which was 45.38% (Cui *et al*., 2020). Ethnicity differences may play a role in contributing to the differences between the findings in this study and other similar studies conducted in different populations. Caucasians may have a higher proportion of normal metabolizers compared to other races/ethnicities though the frequencies are slightly varied depending on the geographical location where the studies were conducted (Zafra-Ceres *et al*., 2013; Hertz *et al*., 2016; Helland *et al*., 2017).

Our results reported *CYP2D6*10* as the most common *CYP2D6* haplotype. Some studies have suggested that this allele increases the risk of breast cancer recurrence for those taking tamoxifen as adjuvant therapy (Kiyotani *et al*., 2008). A study conducted in the Han Chinese population showed that the frequency of *CYP2D6*10* in this population was 45.7% (Lan *et al*., 2018), higher than the frequency of *CYP2D6*10* observed in this study (28.8%). Another important highlight was the relatively high frequency of *\*36* allele observed in this study (0.253) compared to the observed frequency in the PharmGKB database (0.012). Compared to other Asian population, a study conducted in Hong Kong population also recorded a relatively high frequency of *CYP2D6*36* which is 34.1% (Chan *et al*., 2019). Although some *\*36* allele contributed to normal metabolizer status profile, our study observed *\*10/*36* diplotype as the diplotype with highest frequency (0.236), and this diplotype translates as IM phenotype which suggested that *\*36* may play an important role in constructing IM phenotype profiles in Indonesian population. These findings suggested that Indonesian population might be at higher risk of experiencing ineffectiveness of tamoxifen therapy. This was also supported by the high proportion of *CYP2D6* IMs (40.67%) compared to other studies conducted in different populations (Madlensky *et al*., 2011). This was also much higher than the current known global prevalence of IMs which is between 0.4-11% (Gaedigk *et al*., 2017). Even so, some populations also reported a higher proportion of IMs (Hertz *et al*., 2016), suggesting that different populations composed of various ethnicities may play a role in genetic make-up differences of *CYP2D6*. Compared to our result, a similar study conducted in Thailand population showed a relatively high frequency compared to the global prevalence (29.1%), implying that East Asian population may have relatively higher frequency of IM (Sukasem *et al*., 2012). The frequency of NMs observed in this study (54%) was also lower than the current known global prevalence which is between 67-90% (Gaedigk *et al*., 2017).

Different metabolites of tamoxifen and their levels were a predictor of tamoxifen’s efficacy, especially endoxifen levels. Lower endoxifen levels in IMs may indicate lower efficacy of tamoxifen in preventing recurrence (Madlensky *et al*., 2011). Compared to the study conducted by Madlensky *et al*. (2011), the average value of endoxifen levels in IMs observed in this study was higher. The previous study observed the average endoxifen level of IMs to be 8.1 ng/mL while this study recorded an average at 9.6 ng/mL. However, a study conducted in Swedish population found a range of endoxifen level between 2.3-16 ng/mL (Thorén *et al*., 2021), while another study conducted in Singaporean population displayed a range between 1.74–42.8 ng/mL (Lim *et al*., 2011). These suggested that studies conducted with similar interventions but in different populations may find different ranges of metabolite levels.

In this study, we recommended IMs and PMs to adjust their tamoxifen dosage or switch prescription to aromatase inhibitors for patients that were clinically ineligible for consumption of tamoxifen (Goetz *et al*., 2018). We specifically monitored patients who received tamoxifen dose adjustment to 40 mg daily, and our results have shown that participants who received 40 mg of tamoxifen daily all experienced a significant increase across all metabolite levels, including endoxifen levels. This suggested that increasing tamoxifen intake can elevate endoxifen levels as expected and may play a role in increasing the therapeutic effect of tamoxifen. The distribution of endoxifen level in IMs post dose adjustment were similar to the endoxifen level in NMs at the baseline, suggesting that increasing tamoxifen dosage to 40 mg daily for IM participants had successfully let IM participants reach the expected endoxifen levels as observed in NMs.

Gynecological side effects similar to menopausal symptoms such as hot flushes, vaginal dryness, and endometriosis were commonly observed in patients taking tamoxifen (Mourits *et al*., 2001). According to the survey for endocrine symptoms in this study, most participants experienced mild to moderate degree of endocrine symptoms. Despite some of the IM respondents in this study who received dose increase reporting experiencing hot flush, no respondents reported dismissing tamoxifen intake due to the symptom. Hot flush was also commonly reported in patients taking the standard dose of tamoxifen therapy (Kligman & Younus, 2010; Mortimer *et al*., 2008), which means increasing tamoxifen dose does not change side effects of the drug distinctly. Thrombophlebitis, thrombosis, endometriosis, and endometrial cancer were also some of the most concerning side effects of tamoxifen (American Society of Clinical Oncology, 2009; Bergman *et al*., 2000), since they fatally affect patients’ quality of life and life expectancy. None of these side effects were observed in the observed population, but this might also be underestimated due to the short period of follow up on this study. Other studies who have tried to observe tamoxifen side effects occurring in patients with dose increase also concluded that increasing tamoxifen dose did not result in toxicity or short-term increase in side effects (Hertz *et al*., 2016; Dezentjé *et al*., 2015).

These findings concluded that tamoxifen dose adjustment is beneficial enough to increase potential therapeutic effect through the increase of metabolite levels, with no fatal side effects recorded. Although CPIC guideline recommended the first course of action to switch to aromatase inhibitors, our finding demonstrated that tamoxifen dose adjustment is adequate. This is favorable due to: 1) the higher likelihood of potential side effects from aromatase inhibitors than tamoxifen (Garreau *et al*., 2006), 2) lower price of tamoxifen than aromatase inhibitors to allow cost-effectiveness in periodical prescriptions throughout the period of adjuvant therapy.

## Limitations

One of the several limitations of this study was the subjective measurement of side effects using FACT-ES. Due to the subjective nature of answering the survey and rating each symptom, each respondent may have their own perspective on symptom intensity and severity. Another limitation of this study was the relatively short amount of follow up actions observation, which may lead to underestimating the number of certain side effects that may not immediately show up after tamoxifen therapy initiation.

## Conclusion

Our study has shown a considerable proportion of *CYP2D6* intermediate metabolizers (40.67%) in Indonesian women with ER+ breast cancer consuming tamoxifen, suggesting possible ineffectiveness of tamoxifen therapy to prevent recurrence. This was also supported by the significant difference of the endoxifen levels between normal and intermediate metabolizers in our study participants. Dose adjustment of tamoxifen was proven to elevate the level of endoxifen, the metabolite responsible for the anticancer effect of tamoxifen. Increase of tamoxifen intake to 40 mg daily in IM patients did not show any significant or fatal side effects. Given these findings, implementing pharmacogenomic testing of *CYP2D6* on ER+ breast cancer women who are about to undergo adjuvant therapy with tamoxifen may be beneficial to avoid possible inefficacy of tamoxifen in preventing breast cancer recurrence.

## Future Work

Our study has validated the need and benefit of pharmacokinetics and pharmacogenomics analysis, further follow up on the same breast cancer patient cohort is required to better understand the effect and benefits of *CYP2D6* genotyping and endoxifen level measurement towards clinical outcomes such as relapse rate, disease-free survival, and overall survival. Future work may involve monitoring these patients for a longer period of duration when potential side effects were reported previously to appear (Lorizio *et al*., 2012). Monitoring this cohort may be useful to prove the benefit of *CYP2D6* genotyping towards the clinical outcomes of tamoxifen therapy in Indonesian population.

## Data Availability

The data that supports the findings of this study are available as supplementary materials. Any additional data is available from the corresponding author upon reasonable request.

## Acknowledgements

We would like to thank SJH Initiatives and MRCCC Siloam Hospital for their major contribution in patient recruitment, notably Dismas Chaspuri, MD; Arif Winata, MD; and Rachmat Nikijuluw, MD. We would also like to extend our gratitude to The Bioavailability and Bioequivalence Laboratory of Universitas Indonesia, PT Nalagenetik Riset Indonesia, and School of Medicine and Health Sciences, Atma Jaya Catholic University of Indonesia for their support in facilitating sample processing activities. We would like to acknowledge all participants who have voluntarily provided their specimen and time to be involved in this study.

## Authors Contribution

BPM, LLS, AI, YH, H, and SJH contributed to study design. KIJ, MA, and SJH contributed to patient recruitment. BPM and G performed laboratory experiments. KIJ and MA were responsible for patient data management. BPM, KIJ, CM, and AI performed data analysis. KIJ and CM contributed to manuscript writing with supervision from LLS, AI, BPM, and SJH.

## Supplementary

**Supplementary Table 1.** Number of NMs and IMs in each quintile group of endoxifen measured at baseline.

| Quintiles | NMs | IMs | PMs |
| --- | --- | --- | --- |
| Q1 | 8 | 17 | 2 |
| Q2 | 12 | 12 | 0 |
| Q3 | 19 | 11 | 0 |
| Q4 | 15 | 12 | 0 |
| Q5 | 23 | 5 | 0 |

**Supplementary Table 2.**
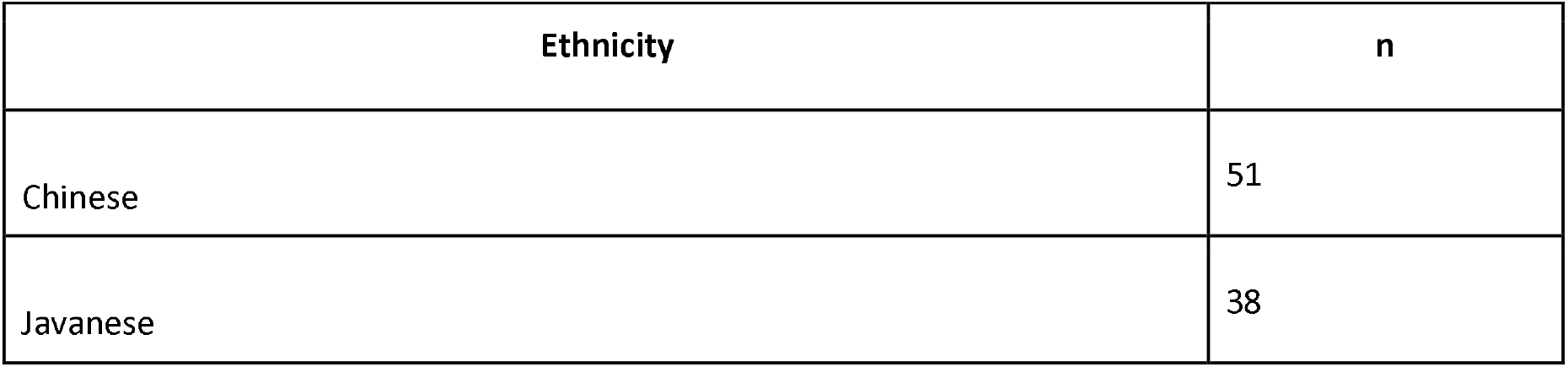
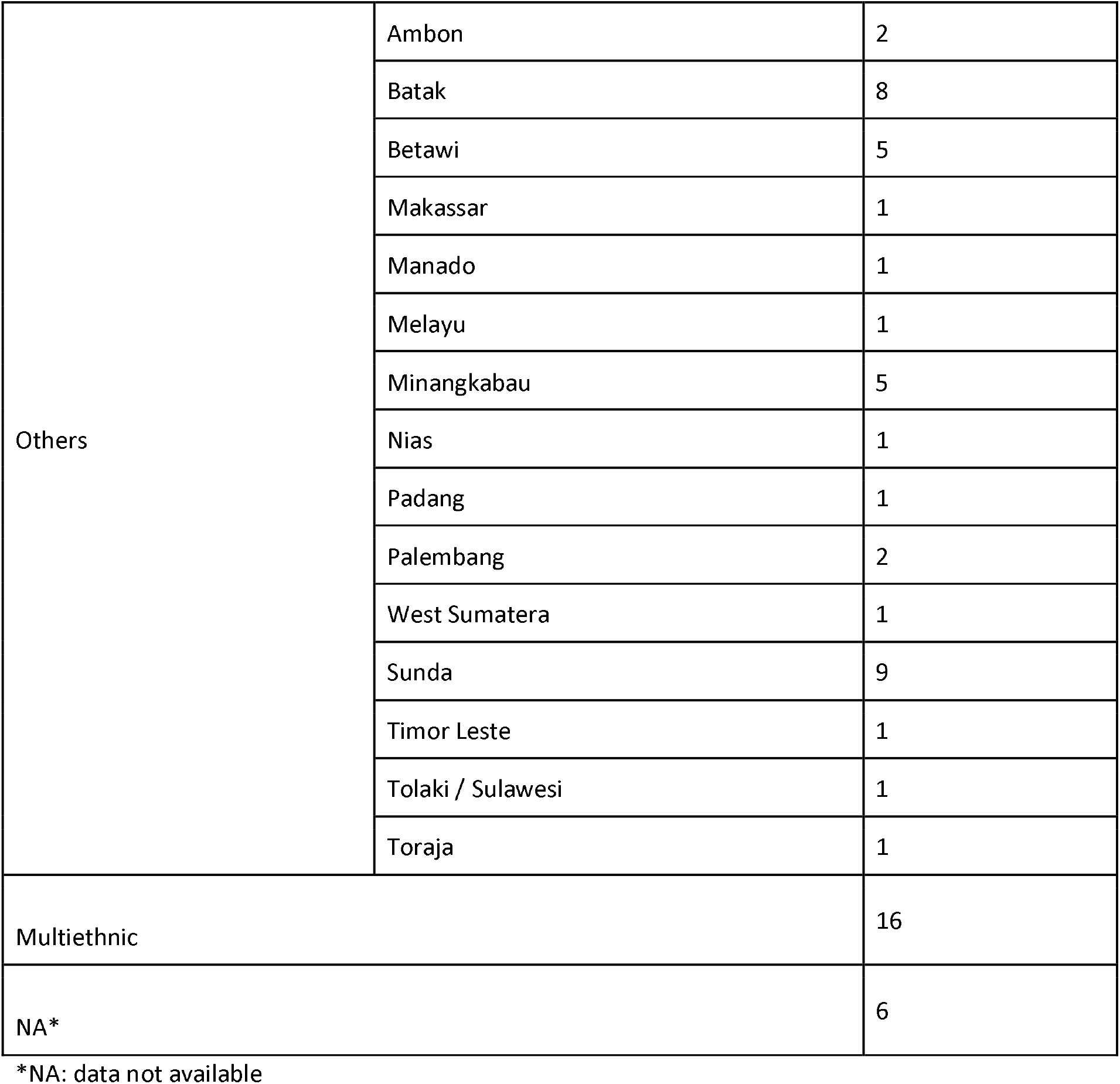
Number of participants in each ethnicity observed. n=150.

| Ethnicity | n |
| --- | --- |
| Chinese | 51 |
| Javanese | 38 |

|  |  |  |
| --- | --- | --- |
| Others | Ambon | 2 |
|  | Batak | 8 |
|  | Betawi | 5 |
|  | Makassar | 1 |
|  | Manado | 1 |
|  | Melayu | 1 |
|  | Minangkabau | 5 |
|  | Nias | 1 |
|  | Padang | 1 |
|  | Palembang | 2 |
|  | West Sumatera | 1 |
|  | Sunda | 9 |
|  | Timor Leste | 1 |
|  | Tolaki / Sulawesi | 1 |
|  | Toraja | 1 |
| Multiethnic |  | 16 |
| NA* |  | 6 |

\*NA: data not available

